# Estimating the generation time for SARS-CoV-2 transmission using household data in the United States, December 2021 – May 2023

**DOI:** 10.1101/2024.10.10.24315246

**Authors:** Louis Yat Hin Chan, Sinead E. Morris, Melissa S. Stockwell, Natalie M. Bowman, Edwin Asturias, Suchitra Rao, Karen Lutrick, Katherine D. Ellingson, Huong Q. Nguyen, Yvonne Maldonado, Son H. McLaren, Ellen Sano, Jessica E. Biddle, Sarah E. Smith-Jeffcoat, Matthew Biggerstaff, Melissa A. Rolfes, H. Keipp Talbot, Carlos G. Grijalva, Rebecca K. Borchering, Alexandra M. Mellis, RVTN-Sentinel Study Group

## Abstract

**Background:** Generation time, representing the interval between infection events in primary and secondary cases, is important for understanding disease transmission dynamics including predicting the effective reproduction number (Rt), which informs public health decisions. While previous estimates of SARS-CoV-2 generation times have been reported for early Omicron variants, there is a lack of data for subsequent sub-variants, such as XBB.

**Methods:** We estimated SARS-CoV-2 generation times using data from the Respiratory Virus Transmission Network – Sentinel (RVTN-S) household transmission study conducted across seven U.S. sites from December 2021 to May 2023. The study spanned three Omicron sub-periods dominated by the sub-variants BA.1/2, BA.4/5, and XBB. We employed a Susceptible-Exposed-Infectious-Recovered (SEIR) model with a Bayesian data augmentation method that imputes unobserved infection times of cases to estimate the generation time.

**Findings:** The estimated mean generation time for the overall Omicron period was 3.5 days (95% credible interval, CrI: 3.3-3.7). During the sub-periods, the estimated mean generation times were 3.8 days (95% CrI: 3.4-4.2) for BA.1/2, 3.5 days (95% CrI: 3.3-3.8) for BA.4/5, and 3.5 days (95% CrI: 3.1-3.9) for XBB.

**Interpretation:** Our study provides estimates of generation times for the Omicron variant, including the sub-variants BA.1/2, BA.4/5, and XBB. These up-to-date estimates specifically address the gap in knowledge regarding these sub-variants and are consistent with earlier studies. They enhance our understanding of SARS-CoV-2 transmission dynamics by aiding in the prediction of Rt, offering insights for improving COVID-19 modeling and public health strategies.

**Funding:** Centers for Disease Control and Prevention, and National Center for Advancing Translational Sciences.

## Introduction

The generation time is a fundamental epidemiological concept for understanding infectious disease transmission dynamics and represents the time between beginning of infection events in primary and secondary cases. Accurate generation time estimation is important for predicting the effective reproduction number (Rt) [1, 2], which helps inform situational awareness and public health decisions.

Previous estimates of SARS-CoV-2 generation times [3, 4, 5, 6] have been reported using data through the early SARS-CoV-2 Omicron variant period (e.g., sub-variant BA.1). However, there is a lack of estimates for more recent SARS-CoV-2 Omicron variant periods of dominance (e.g., sub-variant XBB), despite evidence showing that generation times have substantially decreased over the course of the pandemic, particularly across the Alpha, Delta, and Omicron variants [7]. This trend has corresponded to substantial increases in population immunity, and the emergence of distinct Omicron sub-variants, which may have impacted the generation time of SARS-CoV-2 [8].

In this study, we adapted a previously published method [9] to estimate SARS-CoV-2 generation times using data from more recent SARS-CoV-2 Omicron variant periods specifically the sub-variants BA.1/2, BA.4/5, and XBB.

## Methods

### Household data

Participants were enrolled in a case-ascertained household transmission study, called the Respiratory Virus Transmission Network – Sentinel (RVTN-S), from seven sites across the U.S., 2021–2023 [10, 11, 12, 13]. Individuals diagnosed with COVID-19 were enrolled, along with their household contacts, within 7 days of the initial illness onset within the household. Participants were then monitored prospectively for 10 days, and during this follow-up period participants reported daily symptoms, including fever (including feeling feverish and chills), cough, sore throat, runny nose, nasal congestion, fatigue (including feeling run-down), wheezing, trouble breathing (including shortness of breath), chest tightness (including chest pain), loss of smell or loss of taste, headache, abdominal pain, diarrhea, vomiting, and muscle or body aches. Daily nasal swabs were also collected and tested for SARS-CoV-2 via RT-PCR.

### Estimating the generation time

We employed a mechanistic Susceptible-Exposed-Infectious-Recovered (SEIR) compartmental model and used a Bayesian Markov chain Monte Carlo (MCMC) approach for parameter estimation and data augmentation, as proposed by Hart et al. [9]. The data augmentation MCMC approach, based on the methodology from a household transmission modeling study by Cauchemez et al. [14], was also applied in previous studies [15, 16]. We imputed symptom onsets and infection times of cases to estimate both intrinsic and realized household generation times. The intrinsic generation time assumes no depletion of susceptible individuals, reflecting infection in the community with an unlimited supply of susceptible individuals. In contrast, the realized household generation time accounts for a gradual reduction in the number of susceptible individuals as people are infected within the household setting.

The SEIR model incorporates three infectious compartments: asymptomatic, pre-symptomatic, and symptomatic stages. After infection, individuals enter a non-infectious exposed phase before progressing to an infectious state through one of two pathways: either remaining asymptomatic or developing symptoms following a pre-symptomatic phase. Consequently, transmission can occur before the onset of symptoms, depending on the length of the incubation period. We assumed the incubation period had a mean of 2.6 days and a standard deviation (SD) of 1.0 days for the Omicron variant [17].We also conducted a sensitivity analysis by assuming a longer incubation period with a mean of 4.1 days and an SD of 2.7 days [3].

We also estimated the overall infectiousness, which describes the expected number of household transmissions generated by a single symptomatic infected primary case.

We compared estimates for each sub-period to the overall Omicron variant period by calculating the overlapping index, which measures posterior distribution similarity [18, 19]. Values near 100% indicate high similarity with minimal differences, while values close to 0% suggest low similarity with significant differences.

The estimation was performed in R (version 4.3.1) with 1,000,000 MCMC iterations, discarding the initial 20% as burn-in and obtaining posterior distributions by thinning every 100 iterations. Each MCMC chain took approximately 48 hours to run on a single core of a high-performance computing cluster. The code for the estimations is available at https://github.com/CDCgov/covid-generation_time-us.

### Ethics statement

This study was reviewed and approved by the IRB at Vanderbilt University Medical Center (see 45 C.F.R. part 46.114; 21 C.F.R. part 56.114).

## Results

### Household data

We enrolled 745 households with a single primary case (i.e., only one individual within a household exhibited symptoms on the earliest onset date) with laboratory-confirmed SARS-CoV-2, along with their 1,334 household contacts, totaling 2,079 individuals including both primary cases and household contacts. The study period spanned from December 2021 to May 2023, encompassing the period when the Omicron variant predominated in the U.S. This period was further categorized into three sub-periods dominated by the sub-variants BA.1/2 (December 18, 2021 – June 17, 2022), BA.4/5 (June 18, 2022 – January 14, 2023), and XBB (January 15, 2023 – May 1, 2023) [20, 21].

Of 2079 participants, all of whom had at least two valid SARS-CoV-2 PCR tests [22], 69.4% tested positive and reported symptoms (symptomatic infected), 8.5% tested positive but never reported symptoms (asymptomatic infected), and 22.1% tested negative (uninfected) regardless of symptoms (Table 1). Over the study period, the proportion who remained uninfected increased from 18.5% during the BA.1/2 sub-period to 29.0% during the XBB sub-period.

**Table 1.** Participant symptom and infection status by SARS-CoV-2 variant periods: All Omicron (December 2021 – May 2023), and sub-variants BA.1/2 (December 18, 2021 – June 17, 2022), BA.4/5 (June 18, 2022 – January 14, 2023), and XBB (January 15, 2023 – May 1, 2023).

| Period | Number of participants (households) | Symptomatic infected % (n/N) | Asymptomatic infected % (n/N) | Uninfected % (n/N) |
| --- | --- | --- | --- | --- |
| All Omicron | 2079 (745) | 69.4% (1443/2079) | 8.5% (177/2079) | 22.1% (459/2079) |
| BA.1/2 | 569 (200) | 72.9% (415/569) | 8.6% (49/569) | 18.5% (105/569) |
| BA.4/5 | 1090 (389) | 69.4% (756/1090) | 9.4% (102/1090) | 21.3% (232/1090) |
| XBB | 420 (156) | 64.8% (272/420) | 6.2% (26/420) | 29.0% (122/420) |

### The generation time for the overall Omicron variant period

For the overall Omicron variant period, we found a mean intrinsic generation time of 3.5 days (95% credible interval, CrI: 3.3-3.7) and a mean realized household generation time of 3.0 days (95% CrI: 2.8-3.1) (Figure 1 and Supplemental Table S1). In the sensitivity analysis, assuming the longer incubation period, we found a mean intrinsic generation time of 3.6 days (95% CrI: 3.4–3.9) and a mean realized household generation time of 2.6 days (95% CrI: 2.3–2.9), indicating that the estimates are insensitive to changes in the assumed incubation period.

**Figure 1.**
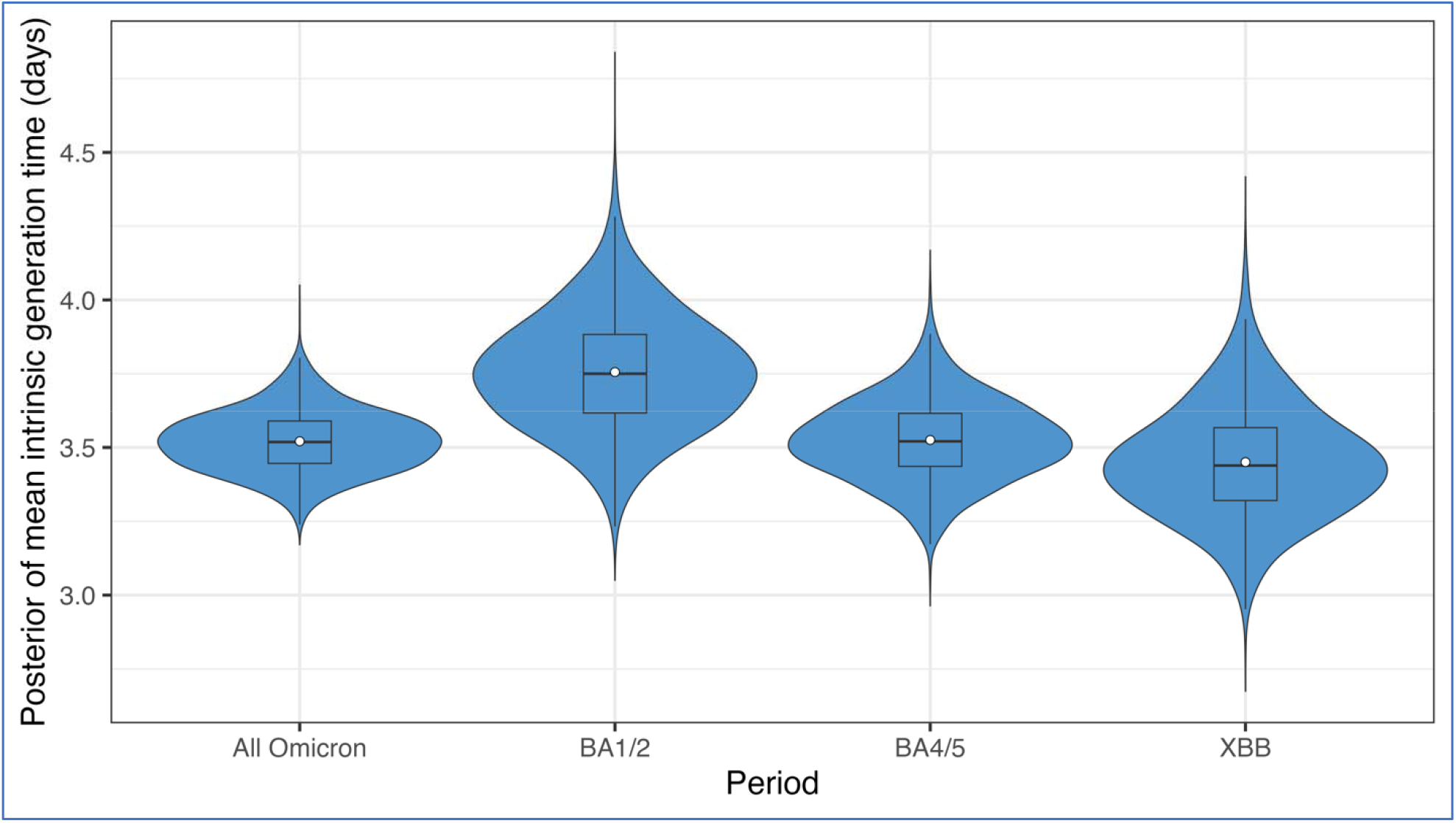
Posterior distribution of mean intrinsic generation time categorized by SARS-CoV-2 variant periods: Omicron (December 2021 – May 2023), and sub-variants BA.1/2 (December 18, 2021 – June 17, 2022), BA.4/5 (June 18, 2022 – January 14, 2023), and XBB (January 15, 2023 – May 1, 2023). The blue violin areas represent the kernel densities. The superimposed box plots show the median values and interquartile ranges. The white dots show the mean values.

### The generation time for the sub-variant periods

The estimates during the sub-periods were similar with more than 40% overlapping to those of the overall Omicron period (Table S1). The mean intrinsic generation time for the BA.1/2 sub-period was slightly longer at 3.8 days (95% CrI: 3.4-4.2). The mean intrinsic generation time for the BA.4/5 and XBB sub-periods was 3.5 days (BA.4/5: 95% CrI: 3.3-3.8; XBB: 95% CrI: 3.1-3.9). The mean realized household generation time for each variant sub-period ranged between 2.9 and 3.1 days (BA.1/2: 3.1 days, 95% CrI: 2.9-3.4; BA.4/5: 2.9 days, 95% CrI: 2.7-3.2; XBB: 3.1 days, 95% CrI: 2.8-3.4).

### The overall infectiousness

The overall infectiousness during the overall Omicron period was 2.5 (95% CrI: 2.4-2.7). This means that, on average, each symptomatic primary case was responsible for approximately 2.5 secondary infections within the household. Additionally, we observed a substantial decrease across each sub-period, starting at 2.8 (95% CrI: 2.5-3.2) during the BA.1/2 sub-period, followed by a slight decline to 2.7 (95% CrI: 2.4-2.9) with 61% overlapping during the BA.4/5 sub-period, and then a significant drop to 2.0 (95% CrI: 1.6-2.3) with only 3% overlapping during the XBB sub-period (Supplemental Figure S1).

## Discussion

Our estimated mean intrinsic generation time for the overall Omicron variant period was 3.5 (95% CrI: 3.3-3.7) days, which is shorter than the estimates for earlier periods of the Delta (4.7 days, 95% CrI: 4.1-5.6), Alpha (5.5 days, 95% CrI: 4.7-6.5) and wild-type variants (5.9 days, 95% CrI: 5.2-7.0) [16, 9]. Our results align with a previous systematic review and meta-analysis [7] that demonstrated a decrease in the generation time (and incubation period) across the Alpha, Delta, and Omicron variants from 2021 to 2023.

This decreasing trend in the generation time may be influenced by a combination of factors. These factors can both shorten the generation time and lower the overall infectiousness, as observed across different Omicron sub-variant periods, partly because fewer transmissions occur in the later stages of the infectious period. These include (1) less infectious infectors due to biological changes in the pathogen, such as specific virological characteristics of the variants/sub-variants, altered viral shedding patterns, or reduced viral load that lowers virus transmissibility; (2) less susceptible contacts due to higher population immunity resulting in fewer successful transmissions; and (3) behavioral changes, such as reduced direct contact with household members and more routine contact with others due to the relaxation of pandemic mitigation measures [8]. However, the exact underlying reasons for this trend remain uncertain.

The shorter generation time suggests transmission occurred earlier, implying faster transmission dynamics, which has implications for public health measures. If it was primarily driven by virologic factors or altered viral shedding patterns, isolation periods might be shortened and focused more intensively on the early days. Importantly, we observed a decrease in the overall infectiousness between the Omicron sub-variant periods. Despite the principle that a shorter generation time does not necessarily correlate with higher overall infectiousness, the combination of a shorter generation time paired with reduced infectiousness suggests that transmission might have occurred faster during the XBB sub-period compared to the earlier Omicron sub-periods, although the overall number of transmission events was lower.

Our intrinsic generation time estimates exceed a previous estimate of 2.9 days [3], which is currently used to predict Rt and to infer the current epidemic growth status for states and territories in the U.S. [1]. This suggests that some current efforts to forecast or model COVID-19 may rely on earlier estimates, which could introduce biases. For example, given historical surveillance data, an underestimated generation time would overestimate Rt and prediction trends. Updating generation time estimates to reflect more recent data and current community transmission settings could improve the accuracy of real-time infection trend predictions [2].

Our estimated realized household generation time for the overall Omicron variant period was 3.0 days (95% CrI: 2.8-3.1), half a day shorter than the intrinsic generation time. Similar estimates were found in earlier studies [3, 4, 5], and a pooled mean for Omicron BA.1 of 3.0 days (95% confidence interval, CI: 2.5-3.5) [7] closely matches our estimates for the BA.1/2 sub-period at 3.1 days (95% CrI: 2.9-3.4). Furthermore, Wang et al. [6] found that the mean realized household generation time of Omicron BA.5 variants was 2.8 days (95% CrI: 2.4-3.5), also closely matching our estimate for the BA.4/5 sub-period at 2.9 days (95% CrI: 2.7-3.2). This consistency confirms the robustness of our estimates.

A limitation of our study is that we estimated the generation time across the entire study population, regardless of prior immunity from vaccination or previous infection. Our objective was to provide more up-to-date estimates that reflect the combined effects of shifting population immunity, behavioral changes, and evolving viral lineages, rather than to estimate specific generation times by immune status. While previous research [16] has shown that fully vaccinated individuals may experience slightly longer generation times, our estimates are intended to capture the broader trends across the population. It is worth noting that study participants may not be representative of the general population, such as correlations due to cluster sampling not being addressed. Another limitation is that we did not consider external infection routes outside the households. Household members might have been infected by the community, leading to an overestimation of the overall infectiousness.

## Conclusions

Based on SARS-CoV-2 household transmission data collected from December 2021 to May 2023 in the U.S., we present updated estimates of the intrinsic and realized household generation time. These estimates offer valuable insights into the transmission dynamics of SARS-CoV-2 within communities and households. Our results are crucial for enhancing COVID-19 modeling and public health strategies and highlight the necessity of ongoing evaluation of transmission patterns for optimal outbreak management.

## Data Availability

The household data are available upon reasonable request and upon completion of required approvals. The R code for estimating generation time is available at https://github.com/CDCgov/covid-generation_time-us.

## Supplementary material

**Table S1.**
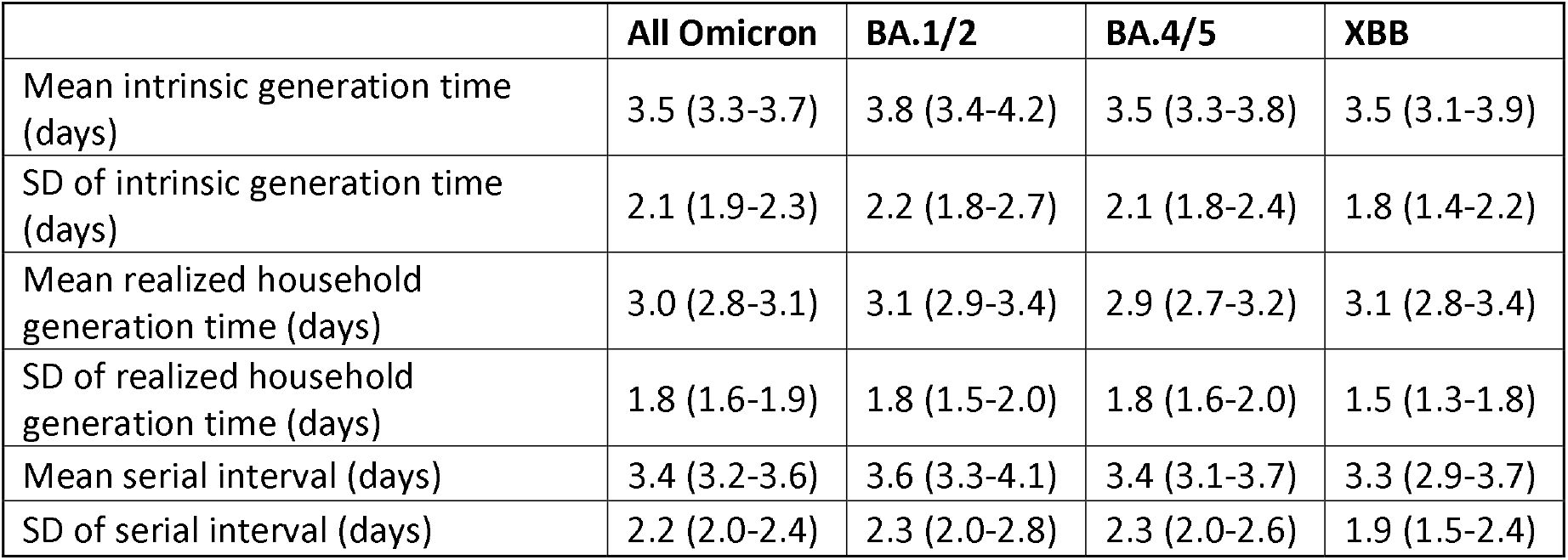
Posterior mean (95% CrIs) of estimates categorized by SARS-CoV-2 variant periods: Omicron (December 2021 – May 2023), and sub-variants BA.1/2 (December 18, 2021 – June 17, 2022), BA.4/5 (June 18, 2022 – January 14, 2023), and XBB (January 15, 2023 – May 1, 2023).

|  | All Omicron | BA.1/2 | BA.4/5 | XBB |
| --- | --- | --- | --- | --- |
| Mean intrinsic generation time (days) | 3.5 (3.3-3.7) | 3.8 (3.4-4.2) | 3.5 (3.3-3.8) | 3.5 (3.1-3.9) |
| SD of intrinsic generation time (days) | 2.1 (1.9-2.3) | 2.2 (1.8-2.7) | 2.1 (1.8-2.4) | 1.8 (1.4-2.2) |
| Mean realized household generation time (days) | 3.0 (2.8-3.1) | 3.1 (2.9-3.4) | 2.9 (2.7-3.2) | 3.1 (2.8-3.4) |
| SD of realized household generation time (days) | 1.8 (1.6-1.9) | 1.8 (1.5-2.0) | 1.8 (1.6-2.0) | 1.5 (1.3-1.8) |
| Mean serial interval (days) | 3.4 (3.2-3.6) | 3.6 (3.3-4.1) | 3.4 (3.1-3.7) | 3.3 (2.9-3.7) |
| SD of serial interval (days) | 2.2 (2.0-2.4) | 2.3 (2.0-2.8) | 2.3 (2.0-2.6) | 1.9 (1.5-2.4) |

**Figure S1.**
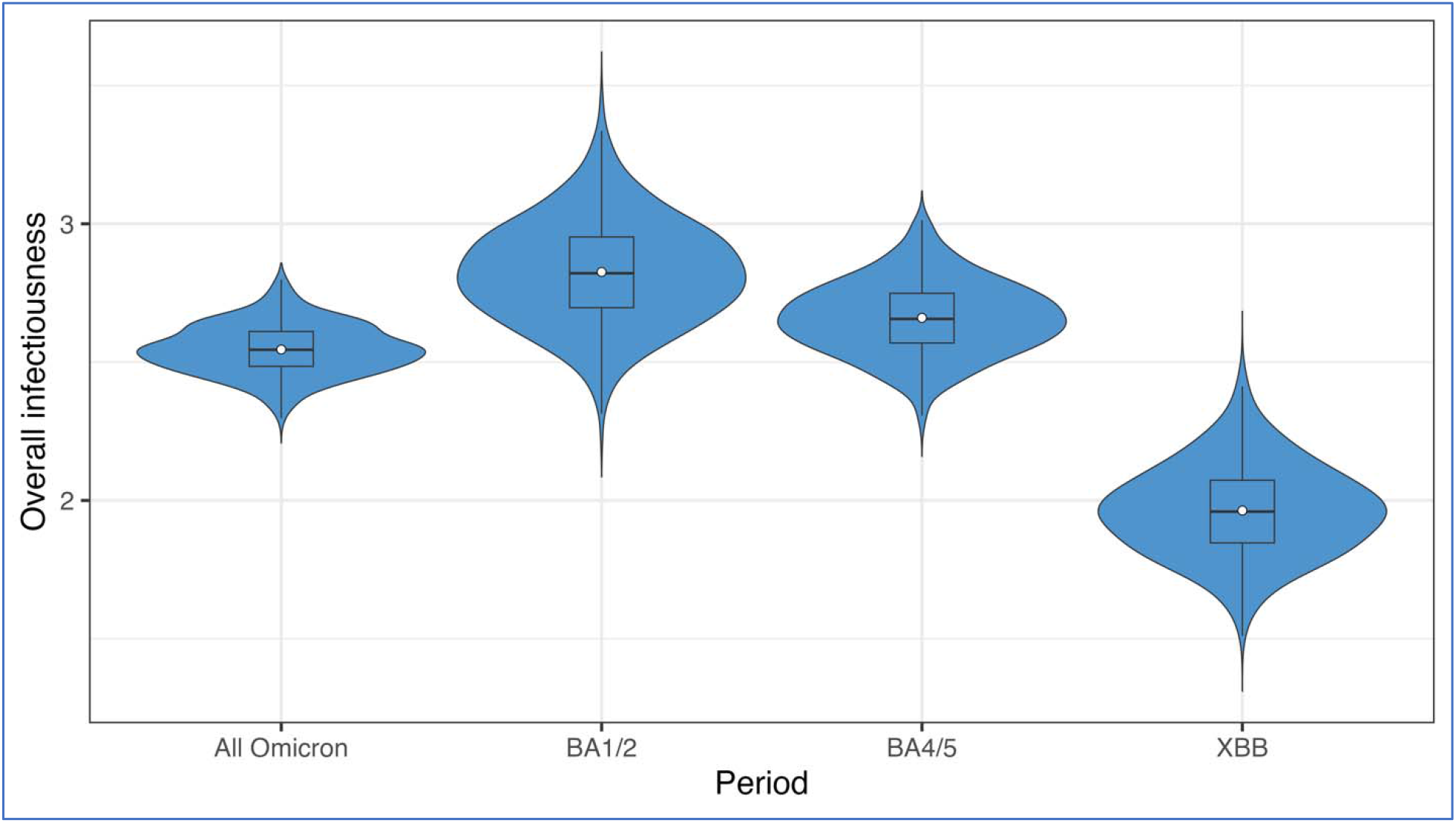
Posterior distribution of overall infectiousness (expected number of household transmissions generated by a single symptomatic infected primary case) categorized by SARS-CoV-2 variant periods: Omicron (December 2021 – May 2023), and sub-variants BA.1/2 (December 18, 2021 – June 17, 2022), BA.4/5 (June 18, 2022 – January 14, 2023), and XBB (January 15, 2023 – May 1, 2023). The blue violin areas represent the kernel densities. The superimposed box plots show the median values and interquartile ranges. The white dots show the mean values.

## Author contributions

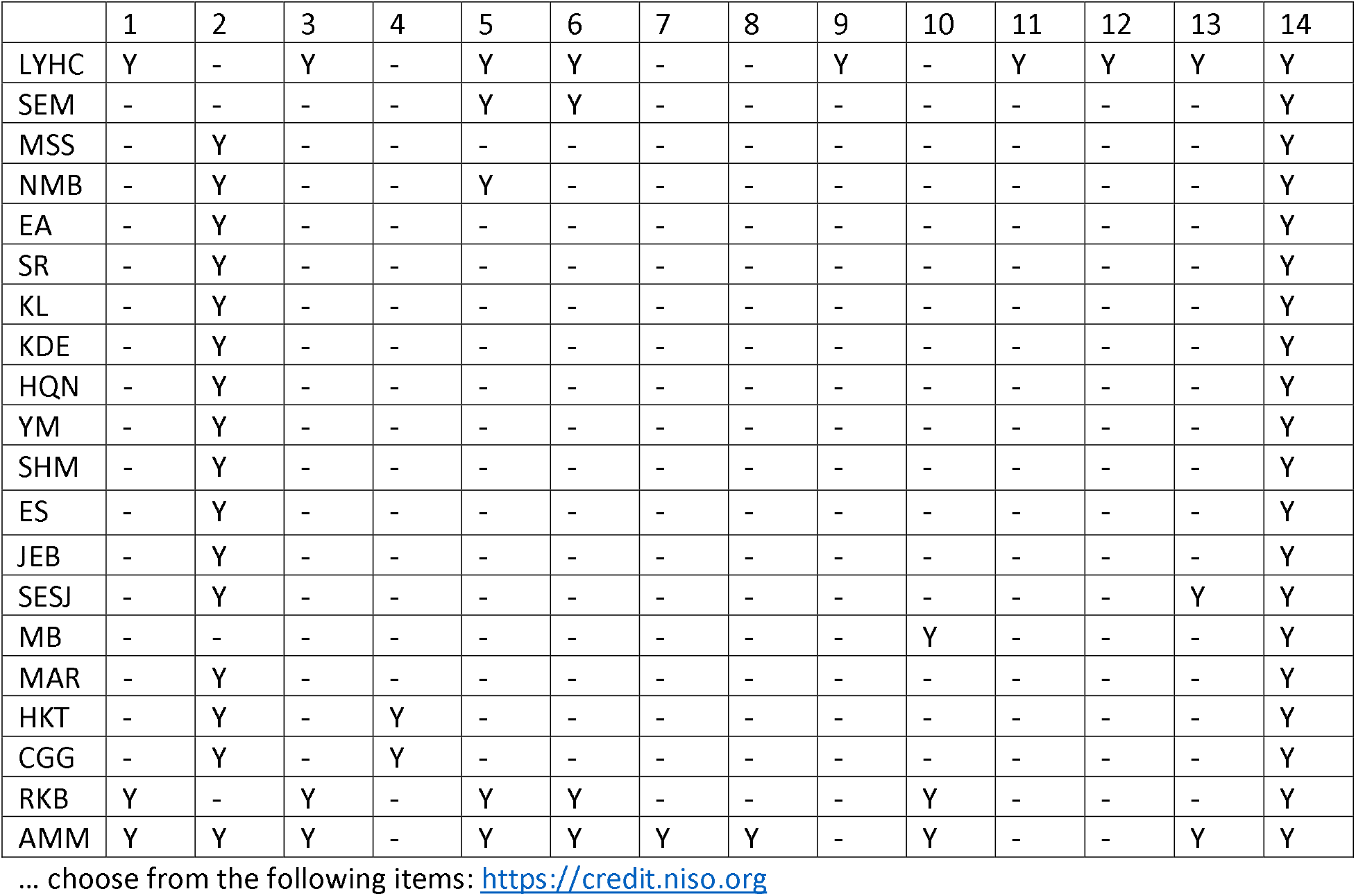

1. Conceptualization

2. Data curation

3. Formal analysis

4. Funding acquisition

5. Investigation

6. Methodology

7. Project administration

8. Resources

9. Software

10. Supervision

11. Validation

12. Visualization

13. Roles/Writing -original draft

14. Writing - review & editing.

## Acknowledgements

- The authors thank the following members of the Respiratory Virus Transmission Network – Sentinel (RVTN-S) study teams:
  a. Vanderbilt University Medical Center: Chris Lindsell, Judy King, John Meghreblian, Samuel Massion, Brittany Creasman, Lauren Milner, Andrea Stafford Hintz, Jorge Celedonio, Ryan Dalforno, Maria Catalina Padilla-Azain, Daniel Chandler, Paige Yates, Brianna Schibley-Laird, Alexis Perry, Ruby Swaimn, Mason Speirs, Erica Anderson, Suryakala Sarilla, Amelia Dodds, Dayton Marchlewski, Timothy Williams, Afan Swan, Onika Abrams, Jackson Resser, Ine Sohn, Cara Lwin, Hsi-nien (Jubilee) Tan, Stephen Yeargin, James Grindstaff, Heather Prigmore, Jessica Lai, Zhouwen Liu, James D. Chappell, Marcia Blair, Rendie E. McHenry, Bryan P. M. Peterson, Lauren J. Ezzell.
  b. Columbia University: Lisa Saiman, Raul A. Silverio Francisco, Anny L. Diaz Perez, Ana M. Valdez de Romero.
  c. Stanford University: Rosita Thiessen, Marcela Lopez, Alondra A. Aguilar, Emma Stainton, Grace K-Y. Tam, Jonathan Altamirano, Leanne X. Chun, Rasika Behl, Samantha A. Ferguson, Yuan J. Carrington, Frank S. Zhou.
  d. Marshfield Clinic Research Institute: Edward A. Belongia, Hannah Berger, Vicki Moon, Gina Burbey, Leila Deering, Brianna Freund, Garrett Heuer, Sarah Kopitzke, Carrie Marcis, Jennifer Meece, Jennifer Moran, DeeAnn Hertel, Joshua Petrie, Miriah Rotar, Carla Rottscheit, Elisha Stefanski, Sandy Strey, Melissa Strupp.
  e. University of Arizona: Ferris Alaa Ramadan, Flavia Maria Nakayima Miiro, Josue Ortiz, Mokenge Ndiva Mongoh.
  f. University of North Carolina: Ayla Bullock, Amy Yang, Quenla Haehnel, Jessica Lin, Julienne Reynolds, Katherine “Katie” Murray, Miriana Moreno Zivanovich, Anna McShea, Brittney Figueroa, Melody Liu.
  g. University of Colorado: Kathleen Grice, Cameron Bendalin, Sonia Chavez, Jolie Granger.
- We also acknowledge the households for their participation.
- L. Y. H. C. thanks the CDC Steven M. Teutsch Prevention Effectiveness (PE) Fellowship.

## Disclaimer

The conclusions, findings, and opinions expressed by authors contributing to this article do not necessarily reflect the official position of the U.S. Department of Health and Human Services, the Public Health Service, the Centers for Disease Control and Prevention, or the authors’ affiliated institutions.

## Declaration of Generative AI and AI-assisted technologies in the writing process

After writing the initial draft and before sending to all coauthors, ChatGPT was used to enhance the clarity and coherence of the writing and to check for grammatical errors. After using this tool, the authors reviewed and edited the content as needed and take full responsibility for the content of the publication.

